# Airborne infection risk in venues with different ventilation strategies – a comparison between experimental, numerical and analytical approaches

**DOI:** 10.1101/2023.06.09.23291132

**Authors:** S. Mareike Geisler, Kevin Lausch, Felix Hehnen, Isabell Schulz, Ulrich Kertzscher, Martin Kriegel, Christian Oliver Paschereit, Sebastian Schimek, Ümit Hasirci, Gerrid Brockmann, Annette Moter, Karolin Senftleben, Stefan Moritz

**Affiliations:** Section of Clinical Infectious Diseases, University Hospital Halle (Saale), Ernst-Grube Str. 40, 06120 Halle (Saale), Germany; Institute of Energy Technology, Department Energy, Comfort and Health in Buildings, Technical University of Berlin, Marchstraße 4, 10587 Berlin, Germany; Biofluid Mechanics Laboratory, Institute of Computer-assisted Cardiovascular Medicine, Deutsches Herzzentrum der Charité, Augustenburger Platz 1, 13353 Berlin, Germany; Charité – Universitätsmedizin Berlin, corporate member of Freie Universität Berlin and Humboldt-Universität zu Berlin, Charitéplatz 1, 10117 Berlin, Germany; Institute of Fluid Dynamics and Technical Acoustics, Hermann-Föttinger-Institute, Chair of Fluid Dynamics, Technical University of Berlin, Müller-Breslau-Str. 8, 10623 Berlin, Germany; Charité – Universitätsmedizin Berlin, Institute of Microbiology, Infectious Diseases and Immunology, Hindenburgdamm 30, 12203 Berlin, Germany

**Author notes:** Correspondence address: University Hospital Halle (Saale), Section of Clinical Infectious Diseases, S. Mareike Geisler Ernst-Grube-Str. 40 06120 Halle (Saale) Germany. These authors contributed equally to this work and share first authorship (S. Mareike Geisler, Kevin Lausch).

## Abstract

The COVID-19 pandemic demonstrated that reliable risk assessment of venues is still challenging and resulted in the indiscriminate closure of many venues worldwide. Therefore, this study used an experimental, numerical and analytical approach to investigate the airborne transmission risk potential of differently ventilated, sized and shaped venues. The data were used to assess the effect size of different mitigation measures and to develop recommendations.

In general, positions in the near field of an emission source were at high risk in all ventilation systems studied, while the risk of infection from positions in the far field varied depending on the ventilation strategy. Occupancy rate, airflow rate, residence time, SARS-CoV-2 virus variants, a high activity level and face masks affected the individual and total infection risk in all venues. The total infection risk was lowest for the displacement ventilation case and highest for the naturally ventilated venue. Therefore, in our study, a properly designed displacement ventilation system is the most effective ventilation strategy to keep airborne transmission and the number of secondary cases low, compared to mixing or natural ventilation.

## Introduction

The severe acute respiratory syndrome coronavirus type 2 (SARS-CoV-2) is the causative agent of the coronavirus disease 2019 (COVID-19) and is transmitted primarily by infectious respiratory droplets and aerosols and less frequently through direct contact or fomites^1–2^. During the COVID-19 pandemic, venues around the world were closed to contain the spread of SARS-CoV-2^3^. Both large- and small-scale events were assumed to increase the risk of virus transmission and thus amplifying the burden of the pandemic. In fact, there are many reports of transmission events in confined and poorly ventilated indoor spaces, partly due to airborne aerosols^4–5^. However, recent studies have shown that the event-related risk of contracting SARS-CoV-2 was low with a well-functioning ventilation system and an appropriate hygiene concept^6–9^.

Ventilation strategies in venues are very heterogeneous and include a variety of displacement (DV), mixing (MV) and natural (NV) ventilation concepts. The room-specific airflow and consequently the accumulation of airborne pathogens is strongly influenced by the ventilation strategy due to differences in the way of air supply and exhaust^10–11^. In DV systems, conditioned air is supplied at low velocity above the floor directly to the occupied zone, rises due to buoyancy effects and is exhausted at the ceiling. MV systems introduce air at high velocity from the ceiling or side wall outside the occupied zone, to mix with the indoor air, and thus dilute contaminants, and then exhausted. Unlike mechanically ventilated rooms, NV systems use only natural forces such as wind or buoyancy effects to create air movement and to supply fresh air. There are a lot of studies, which reported that DV systems are considered to have a lower risk of airborne disease transmission than MV or NV systems due to the higher ventilation effectiveness and index^10,12-17^. Other authors, however, have reported contradictory results^18–19^, but highlighted the need for a sufficient ventilation rate of ≥3 air changes per ≥ hour (ACH) to effectively reduce the risk of infection with DV^19–20^. Venues are usually complex spaces with multiple areas that require special ventilation concepts to ensure good air quality and a low risk of infection throughout the venue. In the past, however, mechanical ventilation systems of venues was given a low priority in the prevention of airborne diseases, as the focus was primarily on the requirements for quiet operation, thermal comfort and economical energy consumption^21–25^. Although venue studies on the risk of airborne disease transmission have increased since the COVID-19 pandemic, a comprehensive risk assessment comparing and classifying different ventilation concepts with regard to their risk of transmitting infectious aerosols is still lacking. The only large-scale monitoring study was conducted as part of the Events Research Programme (ERP) of the UK Government, analysing the ventilation effectiveness in up to 10 differently sized and ventilated theatres during 90 regular events with spectators using CO_2_ sensors^26–27^. The authors of the study identified poorly ventilated areas despite adequate ventilation rates. However, the lack of controlled study conditions, as well as the general inability of CO_2_ approaches to account for the effectiveness of face masks, air purifiers and the infectivity of individuals, e.g. high emitters, indicated that further research is needed^28^. Few studies examined SARS-CoV-2 transmission via aerosols in single venues using analytical^29–30^, computational fluid dynamic (CFD)^6^ or experimental models^31–32^. The analytical approach, such as the Wells-Riley or dose-response approach, assumes, i.a., that aerosols are instantaneously and uniformly distributed in space^33^. Consequently, the spatio-temporal distribution of aerosols is neglected, resulting in the same risk of infection for every person in the room, regardless of their position. CFD analysis can overcome this problem by simulating and visualising venue-specific aerosol distribution patterns, thus enabling the calculation of individual infection risks, as recently done by several published CFD based studies^11,34-36^. Limitations of this approach are the simplified assumptions of stationary airflow patterns, boundary conditions and ideal airborne particles. Therefore, experimental measurements are needed for the validation of the numerical data and vice versa. Current methodologies use optical systems, CO_2_, tracer gas, artificial aerosols or virus surrogates to investigate infectious aerosol distribution in venues^26,31-32,37-40^. However, direct, fast and easy measurement of sputum-like aerosol particles in the immediate vicinity of the emission source and at various far-field positions in everyday environments has proven difficult. The Aerosol Transmission Measurement System (ATMoS) fills the gap, as it can easily quantify aerosol and droplet transmission between dummies in real time and with high resolution at different environmental positions, even over large distances^41^. ATMoS enables room aerosol distribution and exposure measurements making it suitable for the assessment of various indoor scenarios like different ventilation settings and mitigation strategies^42^.

To the best of our knowledge, this is the first large-scale approach that has used a combination of experimental, numerical and analytical investigations to assess the airborne transmission risk potential of venues with different ventilation strategies. Therefore, ATMoS and CFD analyses were used to assess the airborne transmission risk experimentally and numerically in venues with displacement ventilation (DVV), mixing ventilation (MVV) or natural ventilation (NVV). For this purpose, different emission positions and modes as well as boundary conditions (e.g. occupancy, air flow rate) were taken into account for the risk analyses. The experimentally and numerically derived venue-specific infection risk was then compared with the classical analytically Wells-Riley approach^30^. In addition, the effects of mitigation measures and varying boundary conditions on the risk of infection were investigated by CFD. The experimental measurement setup and the venue-specific data on aerosol amounts are presented in Schulz & Hehnen et al.^42^, while the focus of this manuscript is on the calculation of venue-specific infection risks. Consequently, the results were used to identify critical areas and conduct a ventilation-specific risk assessment, followed by a set of venue- and ventilation-specific recommendations to ensure safe events in future.

## Results

### Spatial distribution of individual infection risks in different ventilated venues using CFD analyses

In general, there was a good agreement between ATMoS and CFD derived ppm values for DVV1 and NVV, but worse for MVV1^42^. Using the concentration of infectious quanta of each occupants breathing zone, the venue-specific individual and total risk of infection P_CFD_ and R_CFD_ could be calculated for different settings and emitter positions.

### Infection risk for a sedentary, passive emitter

For DVV1, MVV1 and NVV the exact locations and number of highly exposed positions varied with the position of the emitter. The CFD simulation of DVV1 with an ascending spectator area showed a directional aerosol distribution with a pronounced aerosol plume behind the emitter for the configuration E6 and B9 (Figure 1A 2+3, Fig. S3A). To this effect, the seats behind the emitter were the most exposed, even with increasing distance, while the positions in front of and next to the emitter remained almost unaffected reflected in low individual infection risks (P_CFD_). The number of people exceeding the acceptable risk R_acc_ threshold of 10^−2^ ranged from 0 (0%) to 11 (11%) for the different emitter positions. For MVV1, a preferential but less directional flow of aerosol particles backwards towards the upper right corner was identified (Figure 1B1-3, Fig. S3B). Positions with increased P_CFD_ were located in all directions around the emitter and resulted in 5 (5%) to 10 (10%) spectators reaching R_acc_ above 10^−2^ in the near- and far-field of the emitter. NVV with an ascending spectator area showed a directional aerosol distribution with a pronounced aerosol plume behind the emitter, especially for the emitter position A14 (Figure 1C2 + Fig. S3D), similar to DVV1. Seats behind the emitter had the highest P_CFD_ values, even with increasing distance, but unlike DVV1, the emission of aerosol particles resulted in contamination of the entire venue resulting in 22 (9%) to 170 (70%) spectators exceeding R_acc_ above 10^−2^.

**Figure 1:**
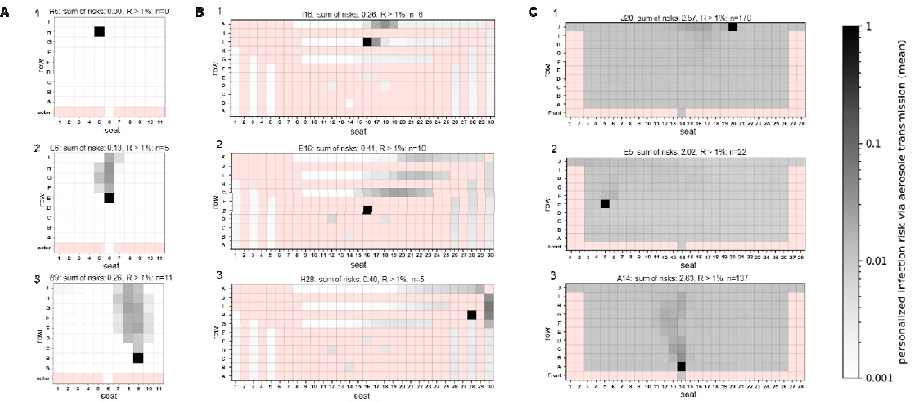
Distribution of the numerical derived individual (P_CFD_) and total risk of infection (R_CFD_) for the venues DVV1, MVV1 and NVV considering a sedentary, passive emitter. Infection risk plots for the venues with displacement ventilation (A), mixing ventilation (B) and natural ventilation (C) are shown. (A) At DVV1, the emitter was located at H5 (1), E6 (2) and B9 (3). (B) MVV emitter positions were at I16 (1), E16 (2) and H28 (3). (C) The positions of the NVV emitters were at J20 (1), A14 (2) and E5 (3). The individual risk of infection is plotted for each position, except for the red positions, as these do not represent seats in the audience. The sum of risks for each venue and emitter positions as well as the number of spectators with R > 1% are indicated above the plots.

The numerically derived risk of airborne transmission R_CFD_, corresponding to the number of new COVID-19 infections, also referred to as secondary cases, varied depending on the position of the emitter and the ventilation strategy. DVV1 showed the lowest R_CFD_ values compared to MVV1 and NVV ranging from 0.00 (H5) to 0.26 (B9) (Figure 1A1-3). For MVV1, the number of new COVID-19 cases was slightly higher, ranging from 0.29 to 0.41 (Figure 1B1-3). R_CFD_ for NVV was about 2 to 2.6 (Figure 1C1-3).

### Infection risk for a high emitter (90^th^ percentile)

In the case of a high emitter, the distribution of aerosol particles and P_CFD_ was similar and dependent on the position of the emitter as for a medium emitter for all ventilation strategies studied (Figure 2). The zone of increased risk was much more pronounced and wider for the DVV1 and NVV cases. In general, an enhanced release of infectious aerosol was associated with an increase in individual and total infection risk at all venues. For DVV1, the number of spectators above R_acc_ remained unchanged for H5 but increased by 2.4 to 2.9-times for E6 and B9, representing 12% to 32% of spectators (Figure 2A1-3). At MVV1 and NVV, a high emitter resulted in the distribution of aerosol particles throughout the venue. This was associated with increased P_CFD_ values at all positions, as demonstrated by almost 100% of spectators achieving R_acc_ above 1% (Figure 2B+C 1-3).

**Figure 2:**
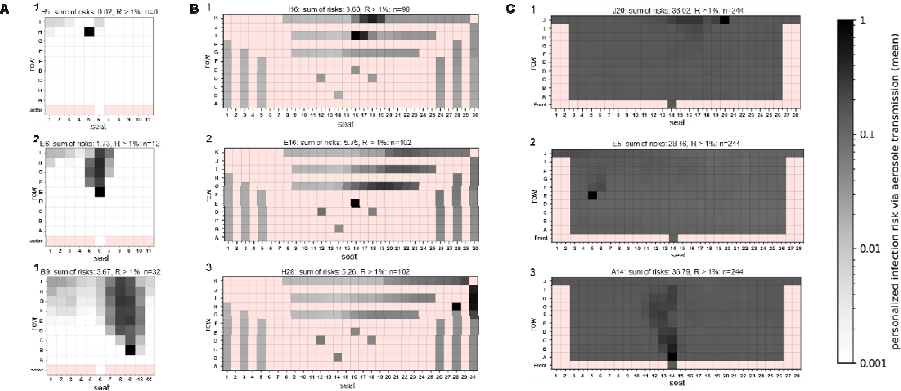
Distribution of the numerical derived individual (P_CFD_) and total risk of infection (R_CFD_) for the venues DVV1, MVV1 and NVV considering an high emitting individual (90^th^ percentile) Infection risk plots for the venue with displacement ventilation (A), mixing ventilation (B) and natural ventilation (C) are shown, considering a high emitter. (A) At DVV1, the emitter was located at H5 (1), E6 (2) and B9 (3). (B) MVV emitter positions were at I15 (1), E16 (2) and H28 (3). (C) The positions of the NVV emitters were at J20 (1), A14 (2) and E5 (3). The individual risk of infection is plotted for each position, except for the red positions, as these do not represent seats in the audience. The sum of risks for each venue and emitter position as well as the number of spectators with R > 1% are indicated above the plots.

R_CFD_ increased by a factor of about 13 to 14 for all emitter positions at all venues (Figure 2A-C). For DVV1 the number of secondary infections was highest for B9 with 3.67 and lowest for H5 with 0.02. R_CFD_ ranged from 3.88 to 5.75 for MVV1 and 28.46 to 36.79 for NVV.

### Comparison of the experimental (R_ATMoS_), numerical (R_CFD_) and analytical (R_analyt_) derived risk of infection for different ventilated venues

The analytical risk of infection R_analyt_ was calculated for all venues and both emission modes according to Peng et al.^30^ and was compared with R_ATMoS_ and R_CFD_ (Table 1). Considering a medium and high emitter at DVV1, R_ATMoS_ and R_CFD_ showed good agreement in 3 out of 4 emitter positions with slight differences, but both were lower than R_analyt_. MVV1 showed comparable values for R_ATMoS_, R_CFD_ and R_analyt_. For the NVV position A14, the values for R_ATMoS_, R_CFD_ and R_analyt_ were heterogeneous, with the lowest value for R_analyt_ for medium and high quanta emission rates. A high emitter led to an overall ∼14-fold increase at all venues and emitter positions for R_ATMoS_, R_CFD_ and R_analyt_.

**Table 1:** Comparison of the experimentally (R_ATMoS_), numerically (R_CFD_) and analytically (R_analyt_) derived total risk of infection. The emitter positions investigated were B9, E6 and H5 in DVV1, E16, H28 and I16 in MVV1 and A14, E5 and J20 in NVV for a silent, sedentary, and high emitter. To obtain R_ATMoS_, the mean value of the seven absorber-specific P_ATMoS_ values of one measurement was calculated and multiplied by the total number of spectators. R_CFD_ represents the sum of the individual infection risks P_CFD_. R_analyt_ was calculated according to Peng et al.^30^ The total risk of infection coincides with the number of new COVID-19 infections and refers to as secondary infections.

| venue | emitter position | sedentary, passive |  |  | high emitter |  |  |
| --- | --- | --- | --- | --- | --- | --- | --- |
| | | $R_{ATMoS}$ | $R_{CFD}$ | $R_{analyt}$ | $R_{ATMoS}$ | $R_{CFD}$ | $R_{analyt}$ |
| DVV1 | B9 | 0.14 | 0.26 | 0.35 | 1.91 | 3.67 | 5.03 |
|  | E2 | 0.14 | 0.12 |  | 1.87 | 1.70 |  |
|  | E6 | 0.19 | 0.13 |  | 2.59 | 1.73 |  |
|  | H5 | 0.04 | 0.00 |  | 0.55 | 0.02 |  |
| MVV1 | E16 | 0.33 | 0.41 | 0.35 | 4.57 | 5.75 | 4.88 |
|  | H28 | 0.27 | 0.40 |  | 3.76 | 5.26 |  |
|  | I16 | 0.23 | 0.26 |  | 3.27 | 3.60 |  |
| NVV | A14 | 1.48 | 2.63 | 0.79 | 20.01 | 36.79 | 11.27 |
|  | E5 | - | 2.02 |  | - | 28.46 |  |
|  | J20 | - | 2.57 |  | - | 36.02 |  |

### Risk evaluation of venues regarding different activity levels, variants of concern and mitigation strategies

The CFD results were used to study the effects of different parameters on the number of COVID-19 secondary cases R_CFD_ compared to R_analyt_ (Table 2). The reference case represented a 2h event with full occupancy and airflow considering the wild-type SARS-CoV2 virus variant and no use of face coverings. Increased activity such as singing or shouting significantly increased R_CFD_ by a factor of 14.4 to 19.3 at all venues compared to a silent, passive emitter. The use of surgical masks reduced R_CFD_ values by a factor of ∼2 to 3 at all venues for both emission profiles. As a result, the number of new COVID-19 cases for a singing or shouting emitter decreased from up to 4.4 to 2.0 for DVV1, 7.9 to 3.4 for MVV1 and 43.7 to 20.5 for NVV but were still about eight times higher than for a silent, passive, non-masked emitter. The use of FFP2 masks reduced the R_CFD_ values obtained with surgical masks by a factor of 7 to 10. Reducing the event duration to 1h decreased the number of secondary infections by ∼2 times, but still showed R_CFD_ > 1 for NVV. In contrast, increasing the residence time to 3h resulted in 1.5-fold higher number of new COVID-19 cases. While R_CFD_ for DVV1 and MVV1 remained < 1, NVV showed R_CFD_ values of 3 to 3.8.

**Table 2:**
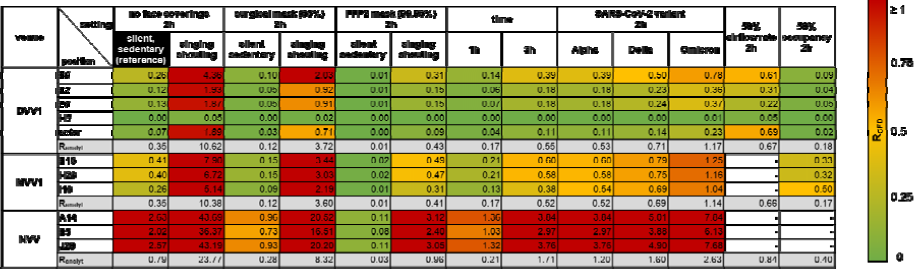
Influence of different parameters and mitigation strategies on the total risk of infection R_CFD_ of venues with different ventilation concepts. The parameter study was performed for different parameters, mitigation strategies, activity levels and emitter positions for the venues DVV1, MVV1 and NVV for the case of a single silent, sedentary emitter (reference). R_CFD_ represents the sum of the individual infection risks P_CFD_ and was calculated for each emitter position of each setting. The R_CFD_ value is a measure of the number of secondary infections. The reference settings (black) were as follows: no face masks, event duration of 2h, SARS-CoV-2 wild-type variant and full occupancy. The efficacy of masks was investigated using surgical masks (65% (0.35) filtration efficiency) and well-fitting FFP2 masks (99.96% filtration efficiency (0.04)). R_CFD_ values were highlighted according to their risk potential using a colour-coded scale with: high risk-R_CFD_ 1 red, medium risk – R_CFD_ = 0.5, low risk – R_CFD_ = 0. Shades of yellow-orange-red and yellow-_≥_green indicate values between the thresholds. R_analyt_ values (dark grey) were calculated according to Peng et al. (2022). Empty boxes indicate the absence of numerical measurements for a given configuration.

Considering the variants of concern (VOC), the number of secondary infections increased 1.5, 2 or 3 times for Alpha, Delta or Omicron. In the case of DVV1, R_CFD_ remained below 1 for all three variants considered. For MVV1, R_CFD_ < 1 was observed for the variants Alpha and Delta, while Omicron resulted in R_CFD_ values of 1.04 to 1.25. Considering NVV, a silent and resting emitter infected with the Omicron variant resulted in 6.13 to 7.8 secondary infections.

The effect of reducing the airflow rate was investigated for DVV1 using CFD analysis, resulting in an increase of 1.7 to 2.3 in airborne infection risk as also seen for R_analyt_.

### Special cases

*Venue with displacement ventilation in the stalls and non-mechanically ventilated balconies (DVV2)* DVV2 contains displacement-ventilated stalls and two naturally ventilated balconies. Numerical (Figure 3, Fig. S3C) and experimental (Fig. S4) measurements observed a pronounced aerosol plume with increased exposure behind the emitter for the position R8S21. Spectators in front of and next to the emitter remained almost unaffected and showed low individual infection risks (Figure 3, Fig. S4). A silent, sedentary emitter placed in the stalls spreaded infectious aerosols up to the balconies. On the contrary, aerosol emissions emanating from the balconies remained their without exposing the stalls but showed a 2.7 times higher risk of infection compared to the stalls (Figure 3A+B, Fig. S3C).

**Figure 3:**
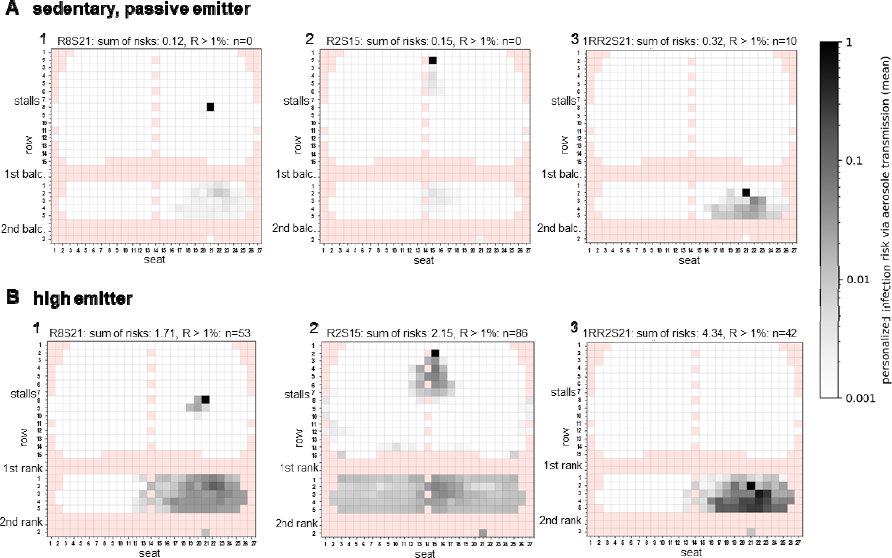
Distribution of the numerical derived individual and total risk of infection for the venue DVV2 with displacement ventilation in the stalls and unventilated balconies. (A, B) Numerically derived infection risk plots for the emitter positions R8S21(1), R2S15 (2) and 1RR2S21 (3) for the silent passive emitter (A) and the high emitter case (90^th^ percentile; B) are shown. The individual risk of infection is plotted for each seat, except for the red shaded positions, as these do not represent seats in the audience. The sum of risk for each venue and emitter position as well as the number of spectators with R > 1% are indicated above the plots. The positions of the stalls, 1^st^ balcony and 2^nd^ balcony are indicated in the graphs. balc. = balcony

In the case of a high emitter, the aerosol partially dispersed over the entire balcony and led to a significant 14.3-fold increase in the airborne infection risk R_CFD_ and R_ATMoS_ (Figure 3A+B3, Fig. S4).

### Infectious actor

In a special configuration, we placed the emitter on stage to experimentally simulate an infectious actor (Figure 4A1). The aerosol that emanated from the infectious actor did not show a directional distribution with a pronounced aerosol plume, as seen for the emitter position B9 (Figure 4A2). A silent, passive actor contributed to low individual infection risks and a low R_CFD_ value of 0.07, which is 3.7 times lower compared to emitter position B9 (Figure 4A1-2). An increased emission and activity rate by a high emitting actor resulted in the exposure of the entire venue, resulting in 1.02 secondary infections and 44 spectators exceeding the critical threshold R_acc_ (Figure 4A3). In contrast, at emitter position B9, mainly spectators behind the emitter were exposed to infectious aerosols, resulting in 3.6-times more secondary infections than in the case of the infectious actor, but showed a lower number of people above R_acc_ (Figure 4A4).

**Figure 4:**
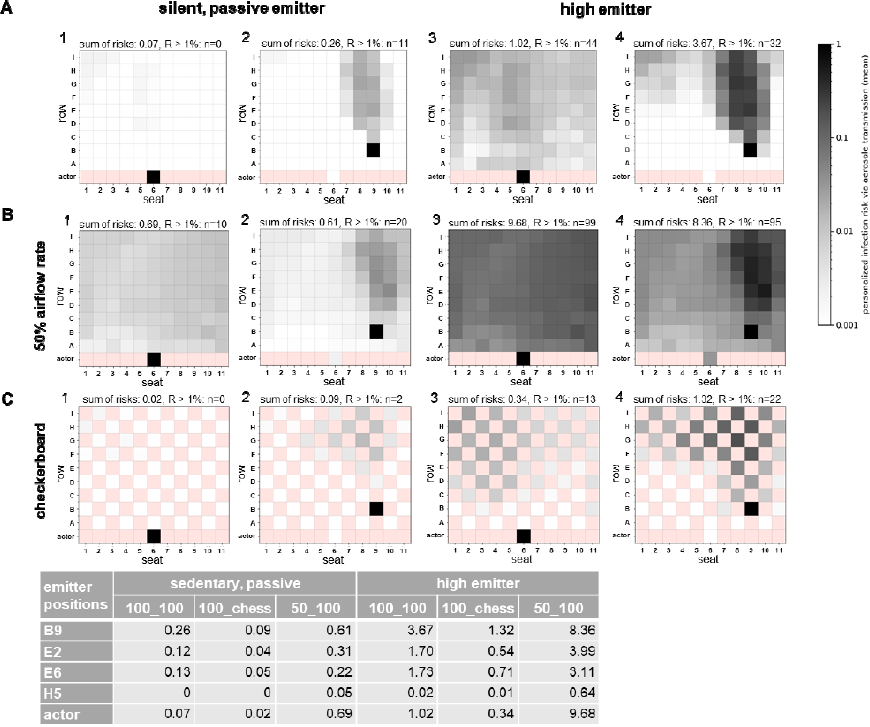
Influence of the emitter stage position (actor), 50% reduced air flow rate and checkerboard pattern seating arrangement on individual (P_CFD_) and total risk of infection (R_CFD_) at DVV1. Infection risk plots for the stage (actor) compared to the audience emitter position B9 for the silent passive emitter (1-2) and the high emitter (90^th^ percentile; 3-4) are shown: (A) standard conditions with full air flow rate (4500 m³/h) and occupancy rate (99 spectators) indicated as 100_100, (B) 50% reduced air flow rate indicated as 50_100 and (C) checkerboard seating arrangement indicated as 100_chess. The individual risk of infection is plotted for each position, except for the red positions as these do not represent seats in the audience. The sum of risk for each venue and emitter position as well as the number of spectators with R > 1% are indicated above the plots and are summarised in the table.

### Variations of boundary conditions

#### Reduced airflow rate

Numerical simulations were performed for DVV1 with a 50% reduced airflow rate (Figure 4B). The pronounced aerosol plume was still observed for emitter position B9, but infectious aerosols were additionally dispersed throughout the venue, resulting in a doubling of spectators exceeding the R_acc_ threshold of 1% and a 2.3-fold increase in secondary infections to 0.61 (Figure 4B2). A silent, passive actor resulted in almost uniform exposure of the entire venue, increasing the total risk of infection by 9.9-fold to 0.69 (Figure 4B1). A high emitting spectator (B9) or actor, combined with a 50% reduction in airflow rate, caused high exposure of the entire venue to infectious aerosols, with almost 100% of spectators exceeding the R_acc_ threshold of 10^−2^ and 8.36 or 9.68 new COVID-19 cases, respectively, representing 8-10% of the total audience (Figure 4B3-4). The high-risk setting with a high emitter in a poorly ventilated venue increased the total risk of infection by 32 (B9) and 138 (actor) times, respectively, compared with the low-risk setting with a silent, passive emitter in a well-ventilated venue.

#### Checkerboard pattern seating

The effect of a 50% reduction in occupancy with checkerboard seating was investigated in venues with displacement (DVV1) and mixing ventilation (MVV1, MVV2) using ATMoS and CFD analyses (Figure 4C, Table 2, Fig. S5-Fig. S8). For DVV1 emitter position B9, the checkerboard pattern seating caused a broadening of the zone of elevated risk, but otherwise showed a qualitatively similar distribution of individual infection risks P_CFD_ for all emitter positions and emission profiles compared with the full seating arrangement (Fig. 4C, Fig. S5). R_CFD_ values of emitter positions investigated in DVV1 were reduced by a factor of 2.6 to 3.5 with checkerboard seating. For MVV1, halving the occupancy rate showed numerically and experimentally a similar distribution of individual infection risks P_CFD_ and P_ATMoS_ (Fig. S6, Fig. S7), but resulted in a different propagation risk pattern for MVV2 compared to full occupancy (Fig. S8). R_CFD_ and R_ATMoS_ values resulted in a decreasing effect of 1.2 to 2 times for MVV1 (Fig. S6, Fig. S7) and 1.3 to 4.5 times for MVV2 (Fig. S7). However, for MVV1 emitter position I16, the R_CFD_ values almost doubled with the checkerboard seating arrangement (Fig. S6).

## Discussion

The COVID-19 pandemic has demonstrated that an appropriate risk assessment is needed to avoid the general and undifferentiated closure of venues in future. To address this shortcoming, venues with different ventilation strategies have been studied experimentally, numerically and analytically in terms of aerosol distribution and exposure to calculate venue-specific infection probabilities and risks compared to the classical analytical approach.

In our study, venues with displacement ventilation and an ascending spectator area posed a low risk of infection under an average emitter scenario with 18.6 quanta h^−1^ (medium emitter scenario), indicated by low individual transmission risks for most spectators and R values well below one. However, the observed pronounced aerosol plume was associated with highly exposed positions in the near- and far-field behind the emitter, while the positions in front of and next to the emitter were almost unaffected, creating characteristic low- and high exposure areas, similar to the results of previous studies^43–45^. The expansion of the aerosol plume and thus the airborne transmission risk is strongly dependent on the position of an infectious person as shown recently^16,44,46^. The radiant wall at the back of the room is assumed to influence the airflow pattern and aerosol dispersion as recently shown^,46^. For MVV1, the airborne transmission risk is less dependent on the position of an infectious individual, but was higher, with a similar number of people exceeding R_acc_ of 10^−2^, compared to DVV1 as shown recently^36,47^. This indicated that infectious aerosols were dispersed throughout MV venues, creating many low- and medium risk positions and a few high-risk positions in the near- and far-field of the emitter, in contrast to DV venues, as also demonstrated for the high emitter scenario. These findings were supported by Makris, Lichtner & Kriegel^43^, which showed that the probability of inhaling aerosol particles at a distance of 1.5 m is twice as high and at a distance of 4 m four times as high for MV cases as for DV cases. Further, MV studies have found high infection probabilities even at longer distances^11,48^. However, predicting highly exposed positions is more difficult as the airflow characteristics in MVV1 are less directional and likely to be sensitive to boundary conditions, as shown by the heterogeneous effects of the checkerboard seating arrangement at MVV1 and MVV2. Moreover, the influence of seasons^49^, air temperatures^50^ and spectator layout^51^ on airflow characteristics has been demonstrated in recent studies on MV, but also on DV cases.

In NVV, the aerosol is distributed in high concentrations throughout the venue, regardless of the position of the infectious source, resulting in the highest airborne transmission risk for each emitter position compared to DV and MV venues, as shown previously^44^. Similarly, recent studies have shown that the risk of airborne transmission does not necessarily decrease with distance in naturally ventilated rooms, as the highest probabilities of infection were observed at longer distances, well beyond physical distance guidelines^11,48^. To keep R < 1 at NVV, the acceptable individual infection risk R_acc_ must be reduced to 0.4%, the number of spectators to 101 (41%) or the exposure time to a maximum of ∼40 min. The maximum number of spectators for a 1.5h event is 133 (55%). Nevertheless, it should be noted that well-designed NV systems are potentially suitable for infection control and provide a cost-effective ventilation approach. However, they are usually highly dependent on natural forces such as wind, open windows or doors and air temperature, and are therefore characterised by unstable and changing airflow patterns, associated with a variety of potential distributions of infection risk in a room^10,12,52-54^.

All high emission scenarios, such as the more infectious SARS-CoV-2 variants, high viral loads or increased activity, were associated with an increased airborne transmission risk. For DVV, this was due to a much more pronounced and wider zone of elevated risks compared to a medium emitter. In contrast, a high emitter in MVV1 distributed the infectious aerosols throughout the venue, resulting in a high risk of infection at any position in the venue, with almost 100% of spectators above R_acc_, comparable to the high emitter scenario in the poorly ventilated DVV1. A high-emitting spectator at NVV resulted in high exposure of the entire venue making NVV a high-risk site with a high potential for super-spreading events, regardless of the position of the infectious person. The maximum residence time or crowding index was significantly reduced to 3.15 min or seven spectators to keep R < 1. Combining the more infectious Omicron variant^55–56^ with a high emitter or singing/ shouting emitter had an additional enhancing effect, resulting in secondary infection rates of 10%, 25% or 50% of the DVV1, MVV1 or NVV audiences. The high emitter case demonstrated greater resilience to different types of emissions for DVV1 compared to the mixing and natural ventilation case. This was confirmed by a study, which showed that even in high emission scenarios, DV performed better than MV systems, which spread the contaminant source over a larger part of the room^57^. To differentiate, high emitting individuals occur only occasionally, but given the substantial super spread potential of SARS-CoV-2, they should be emphasised in the risk assessment to avoid threatening events^58–61^. Furthermore, the case of singing and shouting spectators is less relevant for theatres or operas but considers spectators at concerts and sporting events as well as infectious actors.

FFP2 masks reduced the number of secondary cases by up to 26 times, turning DVV1 and MVV1 into minimal risk sites and reducing the airborne transmission risk of NVV below the critical threshold for pandemic control of one. Similarly, recent studies demonstrated the risk-reducing effect of face masks on COVID-19 transmission^62–65^. However, their limitations became apparent when considering VOCs, virus-rich environments (e.g. hospitals) and prolonged residence in poorly ventilated areas (like NVV), as confirmed by this study and previous research^30,44,64,66^. This highlights the need for a combination of preventive measures. Since the risk of infection increases significantly with the duration of the event^48^, it is recommended to limit the residence time in epidemic settings to a necessary minimum or to consider short breaks^67^. Reducing the number of spectators is also an effective mitigation measure^25,68^. However, experimental and numerical measurements showed a heterogeneous impact of the occupancy rate for the venues with mixing ventilation, similarly to the moderate effects of recent studies^36,68^. For DVV1, the checkerboard seating arrangement resulted in a ∼3-fold reduction in the total risk of infection, clearly indicating an additional downsizing effect beyond the effect of reduced audience size as shown recently^16^.

A reduction in the airflow rate at DVV1 was associated with an up to ∼10-fold increase in the risk of airborne transmission, reaching almost 100% of spectators above R_acc_, supported by a study by Moritz et al.^6^. This showed that the effectiveness of DV is dependent on a room-appropriate and well-adjusted mode of operation as previously demonstrated^19^. This is further demonstrated by DVV2, where a silent passive emitter in the mechanically ventilated stalls resulted in exposure to spectators in the distant unventilated balconies, albeit small, but significantly increased when a high emission scenario was considered, highlighting the need for good ventilation in all areas of the room. Similarly, Adzic et al.^26^ found higher levels of CO_2_, a good proxy for potentially infectious respiratory aerosols^28^, in non-ventilated, but also in ventilated balconies. However, discussions with several stagehands revealed a gap in knowledge about the correct operation and adjustment of the mechanical ventilation system in place and its benefits in reducing airborne disease transmission, resulting in low to moderate airflow rates or partial shutdowns, as also recently discussed^69^. In addition, during the COVID-19 pandemic many venues were operated with 100% outdoor air and with maximum ventilation rates, resulting in high energy consumption and operating costs^26^. Furthermore, it should be emphasised that increasing the flow rate does not necessarily reduce the risk of infection and that close proximity exposure is still likely^46,70-72^. Therefore, ventilation modes and rates need to be optimised to balance the propagation risk reduction with operating system costs.

In summary, ventilation-type specific recommendations for 1, 2 and 3 h events are given in Table 3a+b, considering R well below one. In the case of displacement ventilation case DVV1, the use of surgical masks or a reduction in occupancy is only recommended for events lasting longer than 3h, when Omicron is considered. For MVV1, recommendations for the use of face coverings are given for events of increasing duration, regardless of the infectivity of the virus variant. Reducing the crowding index was not a reliable mitigation measure (Fig. S6). As surgical face masks were sufficient for the wild-type variant, FFP2 masks were suggested for Omicron. The difficulty in predicting the risk of airborne transmission in naturally ventilated venues and the observed high risk of infection justify a general recommendation for FFP2 masks. However, this may not be sufficient for prolonged exposure, particularly to virus variants with increased infectivity, and temporary closure of the naturally ventilated venues should be considered.

**Table 3:** Risk potential and recommendations of risk reduction strategies for venues with different ventilation concepts. (a) The mean values of the venue-specific R_CFD_ values of different emitter positions were calculated for various parameters and mitigation strategies considering the SARS-CoV-2 wild-type and Omicron variant. The total risk of infection coincides with the number of new COVID-19 infections and refers to as secondary infections. In order to keep the threshold for epidemic control R < 1, R_CFD_ values were coloured according to their risk potential: R ≥ 1 red, R = 0.5 yellow, R = 0 green. The values between _≥_ the thresholds are coloured in shades of yellow-orange-red and yellow-green. The pre-pandemic settings with a silent, passive emitter were as follows: no face cover, duration of 2h and full occupancy. (b) Recommendations were given for the conduct of safe events, ensuring R < 0.5 for one to three hour events, considering the SARS-CoV2 wild-type and the Omicron variant for three ventilation concepts. For example, during 3h events in a venue with displacement ventilation and a predominant Omicron variant, an R < 0.5 was achieved by using surgical face masks.

|  | wild-type |  |  | Omicron |  |  |
| --- | --- | --- | --- | --- | --- | --- |
|  | 1h | 2h | 3h | 1h | 2h | 3h |
| **Displacement ventilation** | - | - | - | - | - | surgical face mask<br>reduction in occupancy rate |
| **Mixing ventilation** | - | - | surgical face mask | surgical face mask | surgical face mask | FFP2 mask |
| **Natural ventilation** | surgical face mask | FFP2 mask | FFP2 mask | FFP2 mask | FFP2 mask | FFP2 mask |

The special case of an infectious actor showed that an background actor (silent, passive), posed only a low risk of airborne transmission, which increased significantly when ventilation was halved and the actor became a high emitter with up to ∼9.7 secondary infections, shifting the actor’s emitter position from low risk to the highest risk position with super-spreading potential. In addition, the more realistic scenario of an actor with increased activity, such as singing and shouting, dramatically increased the risk of airborne transmission by 27 times compared to a silent infectious background actor, revealing its threat potential. The risk of infection from actors or singers should be more focused in future in order to minimise the risk to the audience, but also to the ensemble, especially as the stage is often not connected to the ventilation system.

Comparison of the three approaches revealed a good prediction of the overall airborne infection risk by the analytical approach for the mixing ventilation case, indicated by the similarity with numerically and experimentally derived values as shown recently^73–74^. For venues with displacement or natural ventilation, however, the modified Wells-Riley approach over- or underestimated the airborne transmission risk, demonstrating that the strong spatio-temporal dependence of the infection risk could not be captured. The differences between the experimental and the numerical approach are probably due to the random positioning of seven absorbers, while the CFD analysis covers the entire audience. The absorber-specific aerosol concentrations vary strongly depending on their position to the emitter, making it difficult to select a representative set of absorber positions covering the full range of low and high-risk sites. Thus, the calculated venue-representative mean of experimental airborne transmission risk could be biased.

There are some limitations of the study. As ATMoS covered only a few positions at the venues studied, a bias in the experimental results could not be excluded. Validity and reliability of the experimental data could be improved by using more absorbers and by repeated measurements. Furthermore, an event-related R-value threshold of 1 is too high, as an infectious person has additional contacts during the infectious period that must be included to estimate epidemic growth. Concerning the CFD simulations, the assumption of a steady state within the venues is arguable, although the duration of the events is on the order of a few hours. It is likely that during the event the local concentration would increase and converge to the steady state value, which is implied for the risk assessment during the complete event. This shortcoming could be resolved by the unsteady integration of the time-dependent, experienced doses on a stationary flow field or with a fully unsteady simulation approach. Due to the large time frame and the comparatively high temporal resolution, the latter approach would involve rather prohibitively high numerical resources for the given larger venues. Concerning the former approach, numerical experiments were conducted for particular cases, which showed that values close to the steady state concentration were reached within a few minutes for the high-risk regions. Therefore, an additional benefit could not be expected from time-dependent integration results. Additionally, the assumption of constant thermal boundary conditions in a densely occupied event location is critical. The complete knowledge of all environmental boundary conditions and heat load reservoirs would certainly improve the accuracy of the numerical solution. Possibly, the effect would even outweigh the assumption of steady state concentrations. In a similar way, event-specific, relevant boundary conditions (e.g., half-opened doors, reduced ventilation, intensified lighting) had to be disregarded, since they partly depend on personal decision of the responsible technical staff or on the spectator’s behaviour.

Furthermore, the risk models clearly depend on the precise estimation of shed quanta doses and its probability distribution. Due to the lack of a single log-normal distribution which fulfils all given requirements, tests have been conducted where µ was kept constant and σ varied to fit the lower or upper bounds. The effect on the resulting risk was negligible, especially, when compared to the risk differences imposed by a high emitter. However, here again the precise amount of shed quanta, i.e., the chosen quantile of the assumed distribution, is particularly important. On the other hand, this last limitation does not restrict the findings of this study, which result from the comparison of different cases given the same assumed distribution. It rather illustrates the case dependency of super-spreading events.

## Conclusion

Overall, the analytical approach proved to be suitable for the risk assessment of venues with mixing ventilation, although the observed sensitivity to boundary conditions limited its use, even for investigating the effects of different parameters and mitigation strategies. However, to cover various ventilation concepts, and to identify venue-specific high-risk sites and areas of poor air circulation, an individual infection risk assessment through experimental and numerical approaches is required. The results of the study highlighted the wide range of individual infection risks. Low-, medium- and high-risk sites varied according to the ventilation strategy, the emitter position and the emission mode. All three ventilation strategies studied showed high-risk positions in the near field of the emitter, but further distribution in space was different, as shown recently^11,16,25^. At all venues, high-risk positions were also observed well beyond the physical distance guidelines. Venues with displacement ventilation had the lowest overall risk of infection and number of secondary cases with an R_CFD_ value well below one, even when the Omicron variant was considered. The observed directional aerosol distribution allowed prediction of highly exposed positions and the expected number of secondary cases per event. However, in unventilated areas, aerosols can accumulate and locally increase the risk of infection. In venues with mixing or natural ventilation, predicting highly exposed positions is difficult due to the influence of boundary conditions and room parameters (air inlets and outlets, windows, room height, volume) on the room airflow. Face masks provide the best protection against aerosol transmission, but should be combined with other mitigation measures in high risk areas. In terms of pandemic preparedness, the connection of the stage area to the ventilation system should be enforced, as well as raising the awareness of stage technicians and directors of the benefits of a well-adjusted ventilation system in reducing the transmission risk. However, the airflow rate should be balanced between the maximum acceptable individual risk of infection and economically acceptable operating costs.

## Materials and Methods

### Study design

The airborne transmission risk potential of venues with different room characteristics and ventilation concepts was examined using three approaches: experimental measurements using the Aerosol Transmission Measurement System (ATMoS), Computational Fluid Dynamics (CFD) analyses and the analytical Wells-Riley model (Fig. S1). In all three approaches, one infectious person (emitter) was placed in a fully occupied audience. As recent studies have confirmed the presence of people with high viral loads, so-called high emitters^58,61,75,76^, two emission profiles were considered: (I) a sedentary, passive emitter with an average viral load and (II) a slightly active emitter with a high viral load at the 90^th^ percentile. The experimentally and numerically derived absorbed aerosol or quanta concentrations were used to calculate the individual (P) and total risk of infection (R), an analogue of the event reproduction number and an estimate of the effect of a single infectious occupant at an event^69,77^. R also represents the number of secondary infections caused by an infectious individual at an event^78^. Analogous to the basic reproduction number R0, an estimate of the virus transmissibility, R should be kept at < 1 to control disease transmission and epidemic growth^79^. Additionally, special cases were considered by all three approaches: (I) a venue combining displacement ventilation and natural ventilation (DVV2), (II) an infectious actor and (III) varying boundary conditions including reduced airflow rate and checkerboard pattern seating. Furthermore, the effects of mitigation measures, virus variants and varying boundary conditions such as the use of face coverings, residence time, airflow rate and occupancy on the risk of infection were investigated using CFD results. The study design is shown schematically in Fig. S1.

### Venues

To cover commonly installed ventilation systems, venues with displacement ventilation (DVV), mixing ventilation (MVV) and natural ventilation (NVV) were selected. All of the venues studied are theatres with a classic auditorium layout with ascending rows of seat, ranging from 99 to 470. Information on room characteristics and positions of air inlets and outlets are shown in Table S1 and Fig. S2.

### Experimental Measurements

The experimental measurements were carried out with ATMoS and were performed as previously described^41–42^. In brief, ATMoS consists of an emitter that continuously releases a 10%-NaCl-water solution into the environment with a mass flow of 0.43 g/min and an aerosol mean diameter of 2.4 µm with a standard deviation of 1.1 µm. After evaporation, the virus-sized NaCl nuclei remain in the air J and follow the room airflow, thus serving as an ideal virus surrogate. Seven absorbers were distributed in the room, which inhaled the released aerosols at a flow rate of 10 l/min. The absorbed particles were dissolved in ultrapure water and were quantified by conductivity measurement over time. The experimental setup was as follows: 10 min lead-in time to measure the background concentration at each location (no aerosol emission), 27-60 min aerosol emission (Table S1) and 10 min lead-out time (no aerosol emission). For the determination of aerosol emission during regular events, the measurement duration was adapted to event-specific processes, such as the timing of half-times, resulting in different measurement periods. To simulate the influence of body-generated buoyancy effects on aerosol distribution and airflow characteristics, up to 100 heat sources were distributed throughout the venue mimicking a human heat emission of 80 W. In venues with more than 100 seats, experimental measurements were carried out during regular events with spectators and heat sources.

### Calculation of the experimental risk of infection (R_ATMoS_)

Using the inhaled mass of NaCl, the absorber-specific inhaled quanta dose was calculated^39^

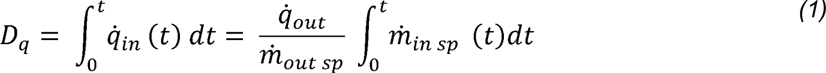

with 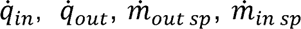, t as quanta input and output rate, mass flow for NaCl output and input and time. According to the Wells-Riley approach, the experimental individual infection risk via aerosols P_ATMoS_ was calculated for each absorber as^80^:

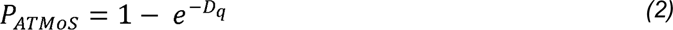

A quanta emission rate of 18.6 quanta h^−1^ was assumed for a sedentary, passive emitter (medium emitter scenario) and 264.68 quanta h^−1^ for a high emitter (high emitter scenario)^30,78^. The distribution of the infection probabilities from the experimental measurements was presented in box plots with median, 0.25 and 0.75 quartile and with minimum and maximum values outside the box. To calculate the venue-specific total infection risk R_ATMoS_, the mean of the seven absorber-specific P_ATMoS_ was calculated and multiplied by the maximum number of occupants (N) per venue as:

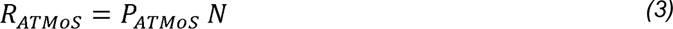

### Computational Fluid Dynamics (CFD)

The presented CFD study includes four venues of particular interest: one displacement ventilation case (DVV1), one multi-purpose mixing ventilation venue (MVV1) and one ascending stage case with nonspecific ventilation concept (natural ventilation, NVV). Furthermore, a special case of displacement ventilation with ventilated stalls and two not-mechanically ventilated balconies (DVV2) was investigated. Numerical analyses were conducted for varying boundary conditions such as occupancy (full vs checkerboard) for DVV1 and MVV1, airflow rates (100% vs. 50%) for DVV1 and emitter positions for DVV1, DVV2, MVV1 and NVV. Using the software Simcenter™ STAR-CCM+ (SIMCENTER), steady state simulations on unstructured finite volume grids are conducted after simplified but detailed reconstruction of the geometric features and boundary conditions based on construction plans, interviews with the responsible technical staff and inspection of the conditions on-site including measurements of the thermal conditions, e.g. temperatures of the environment, the supply air and the surroundings. Flow and energy transport are solved in a segregated manner using the SIMPLE algorithm and the segregated fluid temperature model. Turbulence is modelled by the Realizable k-ε-Modell in a Two-Layer formulation (Wolfstein). Room air is assumed to be a single ε component ideal gas under the influence of gravity. Considering the substantial heat fluxes of lighting, electrical devices and occupants, grey thermal surface-to-surface radiation is applied under usage of view factors. Computer simulated persons (CSP) depict simplified, seated occupants, where the mouth area is specifically distinguished for the insertion of breath tracer gases. CSP are assumed to emit a heat flux of 80 W. Each pre-selected emitter releases a personalized passive scalar with a fictitious, momentum-free mass flux through the mouth area surface cells, which is thereafter transported by convection and diffusion. The passive scalar values throughout the venue’s volume can be referred to their respective source flux and thus local, non-interacting concentrations are obtained within each cell for each emitter. For each CSP a hemisphere of radius 0.23 m (Hemisphere) around the mouth normal vector is defined which acts as a volume-averaged sampling zone assigned to the respective absorbing CSP. The averaged values approximately represent the locally experienced, relative amounts of aerosols shed by the different emitters. These values, along with additional information for further analysis and normalization, are exported and subsequently evaluated in tailored Python scripts. This approach also allows for an a posteriori assignation of the typically uncertain quanta emission rate.

Base sizes of the grid range from 0.1 m to 0.3 m, depending on the size of the venue. Typically, cell sizes are much smaller and rather on the order of a few centimetres in the proximity of CSP, furnishings or equipment. Local refinements, especially on heat or passive scalar emitting surfaces like the CSP, are on the order of millimetres and prism layer cells (4 to 6 layers regularly) support the near-wall solution. Overall mesh sizes range from 3.5 million to 34 million cells. Solution of the flow variables is performed first, while the passive scalar transport equations are solved on the frozen flow field afterwards. Convergence is assumed on the basis of a relative residual drop of at least three orders while simultaneously ensuring constant and physically reasonable monitor values for relevant integral values of the solver variables, e.g., temperature or passive scalar fluxes.

### Calculation of the numerical risk of infection (R_CFD_)

To the present day, there is still uncertainty concerning the quanta emission rate of SARS-CoV-2 aerosols. Peng et al. (2022) established an approximately log-normally distributed emission rate with a mean of 18.6 quanta h^−1^ for a sedentary, passive emitter of the wild-type variant, where the 5th and 95th percentile are located at 8.4 and 48.1 quanta h^−1^, respectively. Buonanno et al.^78^ specify comparable log-normally distributed emission rates for different activity and vocalization levels. Between the two studies the deviations in reported mean values and standard deviations are partially balanced by enhancement factors to compensate for case differentiation. To the authors’ knowledge, there is no log-normal distribution which fulfils all three requirements stated above for the mean and the two given percentiles. Moreover, non-passive behaviour, e.g. (quiet) speaking, is not incorporated in the distribution of Peng et al. (2022). Since the emission profile of occupants is subject to personal variations and behaviour, we assume a combined log-normal distribution with mean value of 18.6 quanta h^−1^ and standard deviation of = 0.720 * ln 10) (≈ 1.65786, where the latter is as suggested by Buonanno et al.^78^. Thus, the distribution to fulfil these conditions is given by LN, (^2^-) where the μ σ desired normal mean is given by μ = 0.672683 * ln(10) ≈ 1.54891.

500.000 pseudo-random number realizations of this distribution have been computed to account for the variability of the quanta emission rate while the mean value of 18.6 quanta h^−1^ was verified. Subsequently, for all realizations and all venues the corresponding quanta doses were calculated based on the locally experienced volume-averaged values within the CSP hemispheres and the event duration according to equation (1) (i.e., steady-state absorption is assumed). By applying equation (2) the local risk P_CFD_ with respect to a given emission rate is evaluated. In a last step, the average of all realizations within a particular venue is calculated as R_CFD_, creating a mapping of the mean, local infection risks with regard to the given emission rate distribution. For further analysis, high emission cases without variation (264.68 quanta h^−1^) are covered. Furthermore, an individual acceptable risk of infection R_acc_ was determined for numerically derived infection risks and set at 10^−2^ in accordance with recent studies^78,81-82^ as the acceptable level of the COVID-19 risk of infection is still unknown. When planning future events, the individual infection risk must be less than R_acc_ to keep the risk of infection to spectators manageable. This enabled the identification of high-risk areas and risk management at the venues studied.

Furthermore, a risk analysis was conducted, considering the effect of mitigation measures, virus variants and varying boundary conditions. In detail, cases with different mask efficiencies (surgical mask: 65%, FFP2 mask: 99.96%), SARS-CoV-2 virus variants (Alpha, Delta and Omicron with enhancement factors as in Table S2) as well as variations of duration (1, 2 and 3 h) and vocalization were compared by taking into account the multipliers of the quanta absorption (see Table S2).

### Modified Wells-Riley approach

The analytical Wells-Riley approach^80,83^ was applied to each venue with modifications^30^ to prove its applicability for a venue-specific infection risk assessment.

### Calculation of the analytical risk of infection (R_analyt_)

For classification of venues in terms of their potential airborne infection risk, the risk parameter H was introduced by Peng et al.^30^. To account for the effectiveness of air distribution of the different ventilation systems and to improve the imperfect well-mixed assumption of the analytical model, the parameter ventilation effectiveness (E_z_) was introduced into the equation of Peng et al.^30^, similar to an approach of Sun & Zhai^68^, by multiplying the air exchange rate (AER) λ by E_z_:

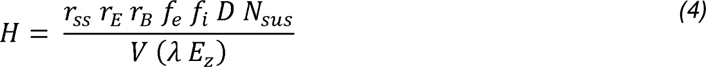

where *r_ss_* is the correction factor for the deviation of average quanta concentration from that of steady state, e.g., for events too short to approximately reach steady state, *r_E_* is the activity-related shedding rate enhancement factor, *r_B_* is the activity-related breathing rate enhancement factor, *f_e_* and *f_i_* are the exhalation and inhalation penetration efficiency for face covering, *D* is the duration of the event, *N_sus_* is the number of susceptible persons and *V* is the indoor environment volume. Ventilation-specific values for E_z_ can be found in the ASHRAE Standard 62.1^15^ and are listed in Table S1 for the venues studied. In considering the worst-case scenario, virus decay and the deposition rate of virus-containing particles in the air were assumed to be low and therefore neglected. The analytically derived total risk of infection (R_analyt_) was calculated with parameter-specific values analogous to Peng et al. (2022) shown in Table S2:

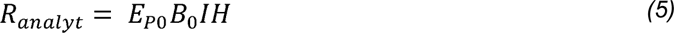

with E_P0_ the SARS-CoV-2 exhalation rate of an resting and only breathing infector, B_0_ the breathing rate of a resting susceptible person and the I the number of infectors present. The breathing rate was set to 0.49 m³/h. The basic configuration represents a typical pre-pandemic event in different venues with the presence of one infectious person and was defined as follows:

- duration of the event: 2h
- occupancy: 100%
- activity level: sedentary, passive
- SARS-CoV2 variant: wild-type
- no face coverings

For surgical masks, a penetration efficiency of 0.35 (65%) was assumed, i.e. 35% of exhaled particles still pass through the mask when both infectious (mask exhalation efficiency 50% (0.5)) and susceptible persons (mask inhalation efficiency 30% (0.7)) wear a mask. A filtration efficiency of 99.96% (0.04) was assumed for a well-fitting FFP2 mask.

## Data Availability

All data produced in the present study are available upon reasonable request to the authors.

## Acknowledgements

We especially thank the venue managers, the stage technicians and the staff of the Puppet theatre Halle, the New Theatre Halle, Opera Halle, the Maxim-Gorki Theatre Berlin and Volksbühne Berlin for the opportunity to carry out the experimental measurements with ATMoS, for their support and for providing floor plans and technical drawings for CFD analyses. We would like to thank Ken J. Lindenberg and Anastasia Strigaleva for their assistance with the CFD analysis and Sophia Wolff and Katharina Schmidt for their assistance with the experimental measurements.

## Author contributions

S.M.G.: conceptualization, methodology, formal analysis, investigation, resources, data curation, writing - original draft, writing - review and editing, supervision, visualization, funding acquisition K.H.L.: conceptualization, methodology, software, validation, formal analysis, investigation, resources, data curation, writing - original draft, writing - review and editing, visualization. F.H.: conceptualization, methodology, validation, formal analysis, investigation, resources, data curation, writing - review and editing, supervision. I.S.: conceptualization, methodology, validation, formal analysis, investigation, resources, data curation, writing - review and editing. U.K.: conceptualization, methodology, resources, writing - review and editing, supervision. M.K.: conceptualization, methodology, resources, writing - review and editing, supervision. O.P.: conceptualization, methodology, resources, supervision, funding acquisition S.S.: conceptualization, methodology, investigation, resources, writing - review and editing. Ü.H.: conceptualization, methodology, formal analysis, investigation, resources. G.B.: methodology, software, formal analysis, investigation, resources, data curation. A.M.: conceptualization, supervision. K.S.: formal analysis, investigation. S.M.: conceptualization, methodology, investigation, resources, writing - review and editing, supervision, project administration, funding acquisition.

## Ethics Statement

The Ethics Committee of the Martin Luther University (Halle, Germany) approved the aerosol measurement experiments with the ATMoS system during regular events with spectators.

