## Supplementary figures and tables for "Airborne infection risk in venues with different ventilation strategies – a comparison between experimental, numerical and analytical approaches"

### Supplements

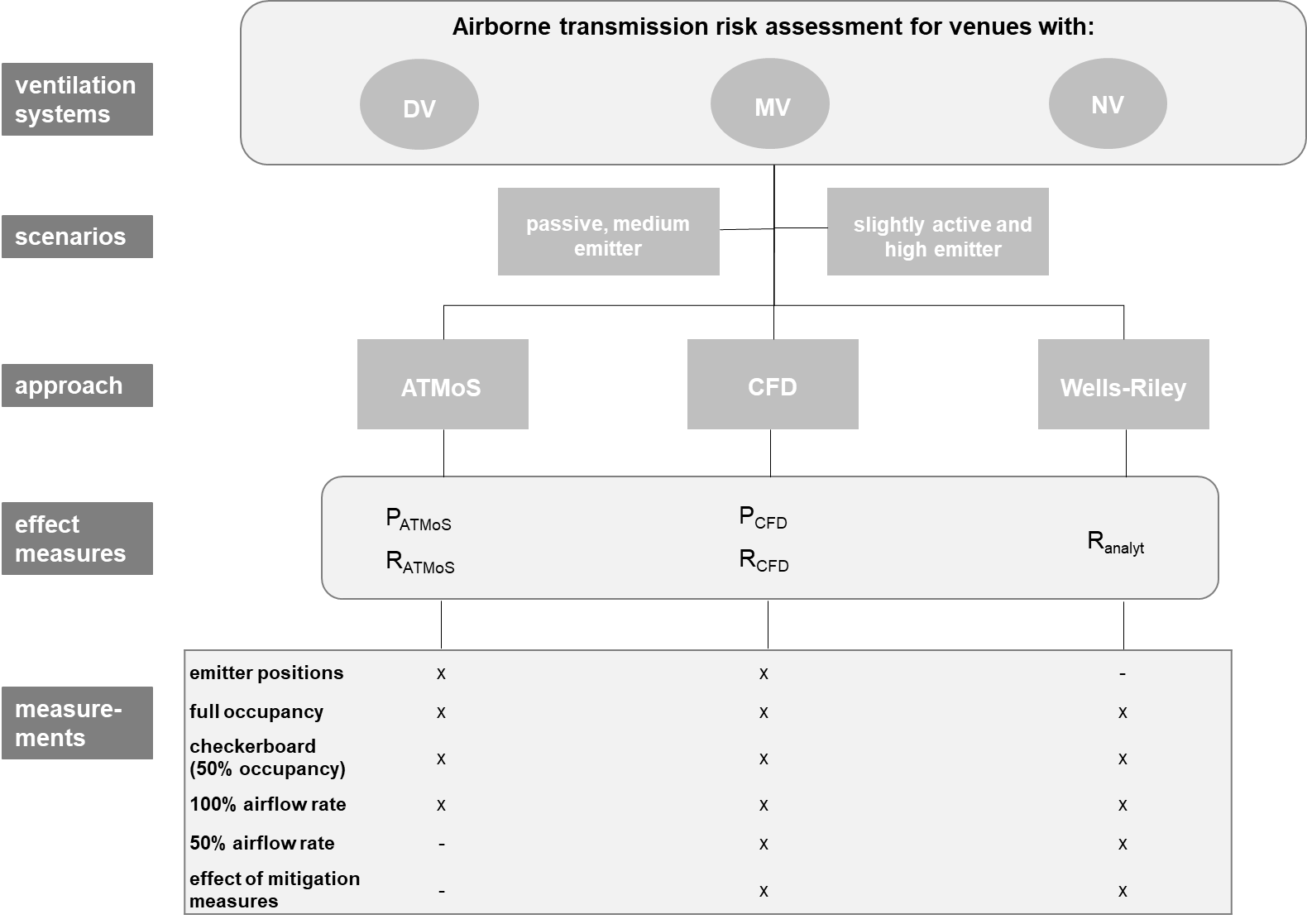

**Fig. S1:	Flow chart of the study design**

Venues with three different ventilation systems, namely displacement ventilation (DV), mixing ventilation (MV) and natural ventilation (NV) were assessed for their experimental (P_ATMoS_, R_ATMoS_), numerical (P_CFD_, R_CFD_) and analytical (P_analyt_) individual and total risk of airborne transmission. Two emission profiles were considered for analyses: a sedentary, passive emitter with an average viral load and a slightly active emitter with a high viral load at the 90th percentile (high emitter). The effect of varying boundary conditions on the risk of infection was investigated, e.g. emitter position, occupancy rate, airflow rate, mitigation measures.

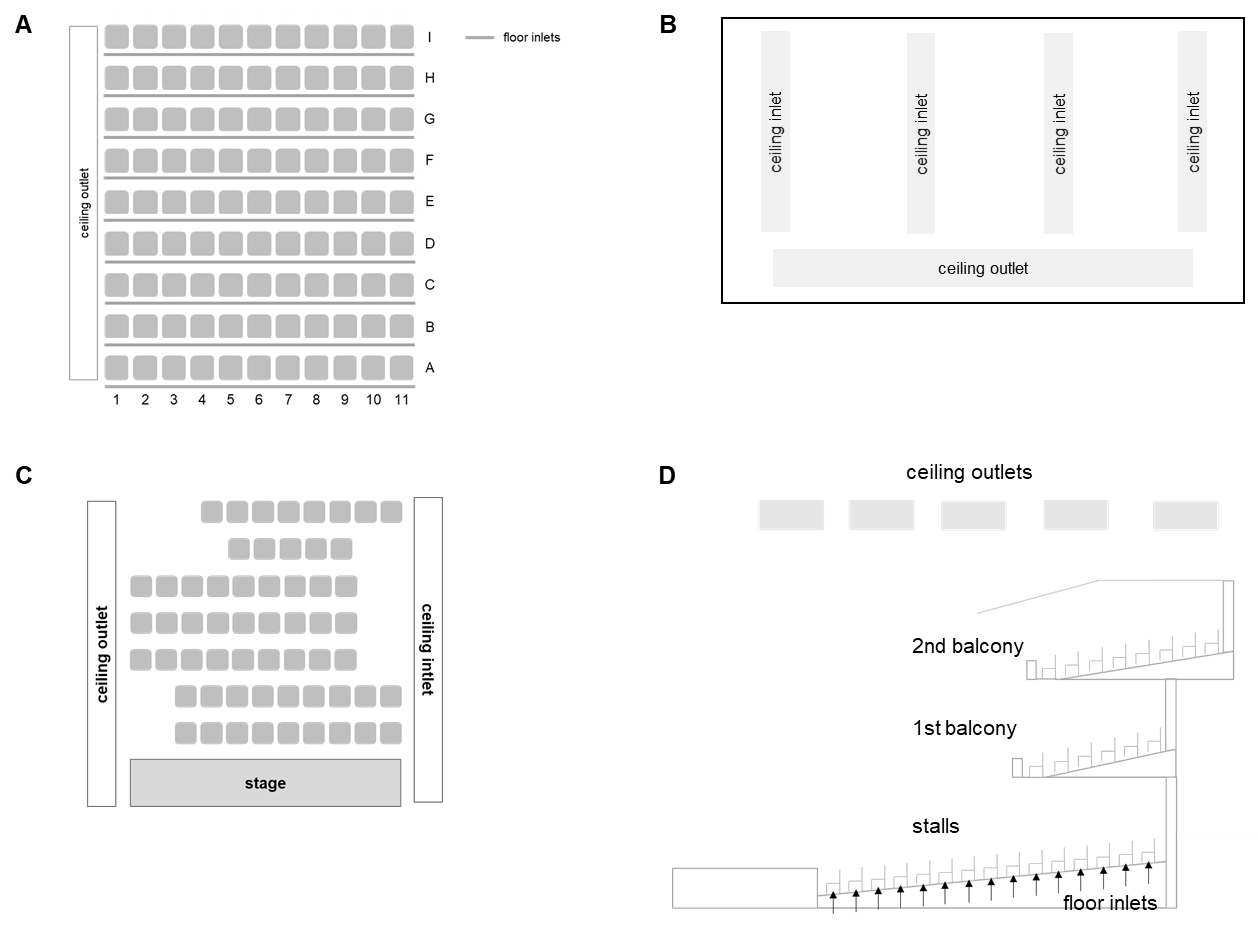

**Fig. S2: Positions of air inlets and outlets of the venues DVV1 (A), MVV1 (B), MVV2 (C) and DVV2 (D)**

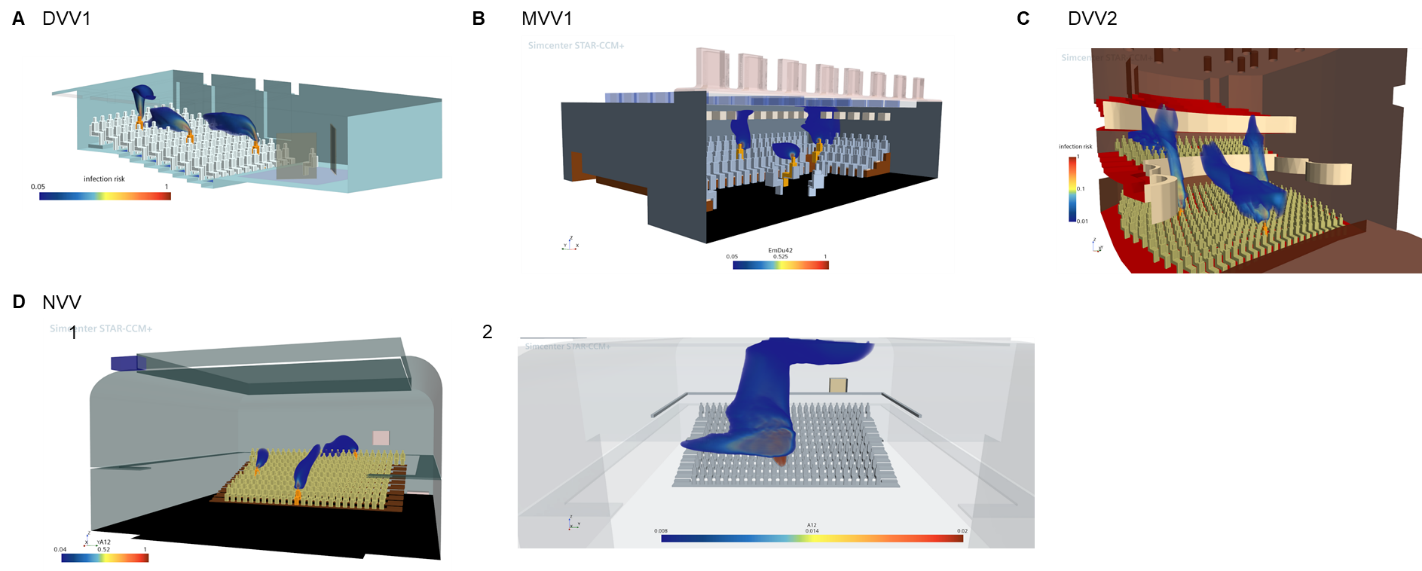

**Fig. S3:	Numerical visualisation of the infection risk or relative contamination for DVV1, MVV1, DVV2 and NVV**

Visualisation of the venue-specific infection risk (A, B, C, D1) and relative contamination level (D2), which relates the local, virtual amount of contaminant to that in the room's exhaust air for better comparability for (A) DVV1 with emitter positions B9, E6 and H5, (B) MVV1 with emitter positions E16, H28 and I16, (C) DVV2 with emitter positions R8S21, R8S7, R2S15 and 1RR2S21 and (D) NVV with emitter position A14, E5 and J20.

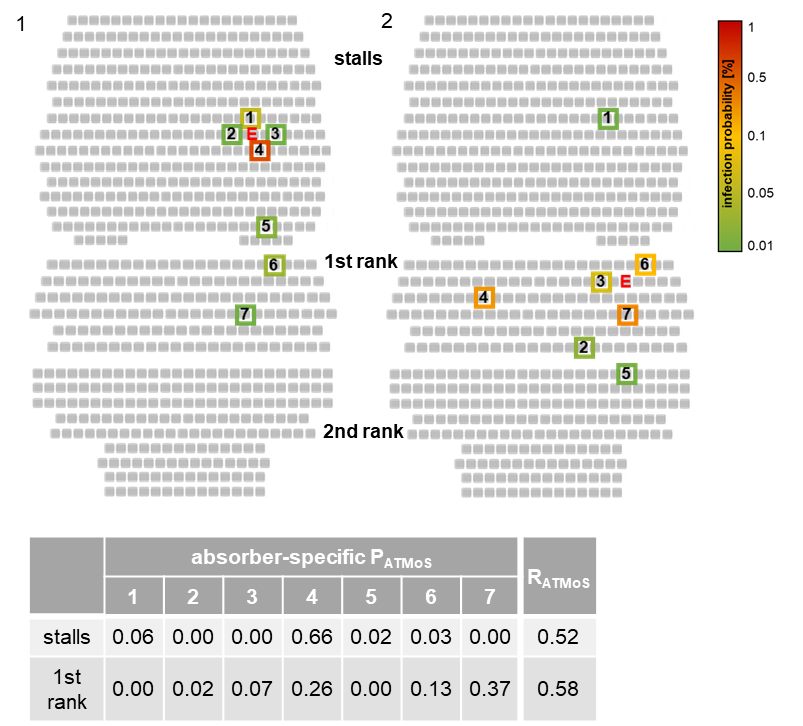

**Fig. S4:**	**Distribution of the experimentally derived individual and total risk of infection for the venue DVV2 with displacement ventilation in the stalls and unventilated balconies**

The experimentally derived absorber-specific individual risk of infection P_ATMoS_ is shown for the emitter position R8S21 in the stalls (1) and for 1RR2S21 in the first rank (2), where “E” and coloured boxes indicate the positions of the emitter and the seven absorbers. The boxes of the absorbers are coloured according to their measured concentration with shades of green, yellow and red, representing low, medium and high measured values. P_ATMoS_ was calculated for each absorber using the absorbed NaCl mass and a quanta emission rate of 18.6 quanta h^-1^. The values for each absorber are summarised in the table. The absorber-specific P_ATMoS_ values were used for the calculation of R_ATMoS_.

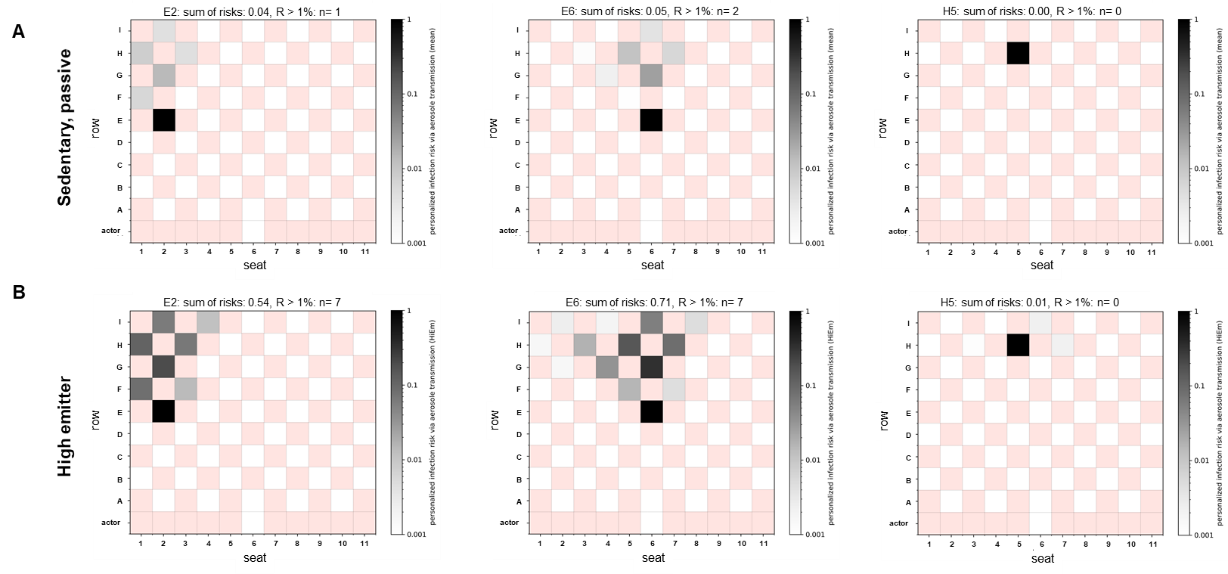

**Fig. S5:	Influence of the checkerboard pattern seating arrangement on individual (P_CFD_) and total risk of infection (R_CFD_) at DVV1**

Infection risk plots for the emitter position E2, E6 and H5 for the silent passive emitter (A) and the high emitter (90^th^ percentile; B) are shown with full air flow rate. The individual risk of infection is plotted for each position, except for the red positions as these do not represent seats in the audience. The sum of risk for each venue and emitter position as well as the number of spectators with R > 1% are indicated above the plots and are summarised in the table.

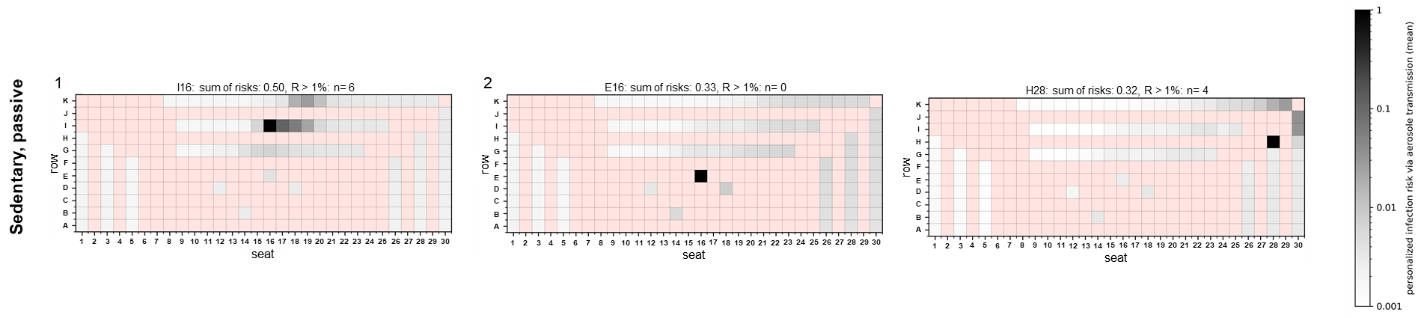

**Fig. S6:**	**Distribution of the numerical individual (P_CFD_) and total risk of infection (R_CFD_) for MVV1 for a silent, passive emitter considering checkerboard seating arrangement**

Infection risk plots for the emitter positions I16 (1), E16 (2) and H28 (3) are shown for a silent, passive emitter in a checkerboard arrangement of spectators. The individual risk of infection is plotted for each spectator, except for the red positions as these do not represent seats in the audience. The sum of risks for each venue and emitter position as well as the number of spectators with R > 1% are indicated above the plots.

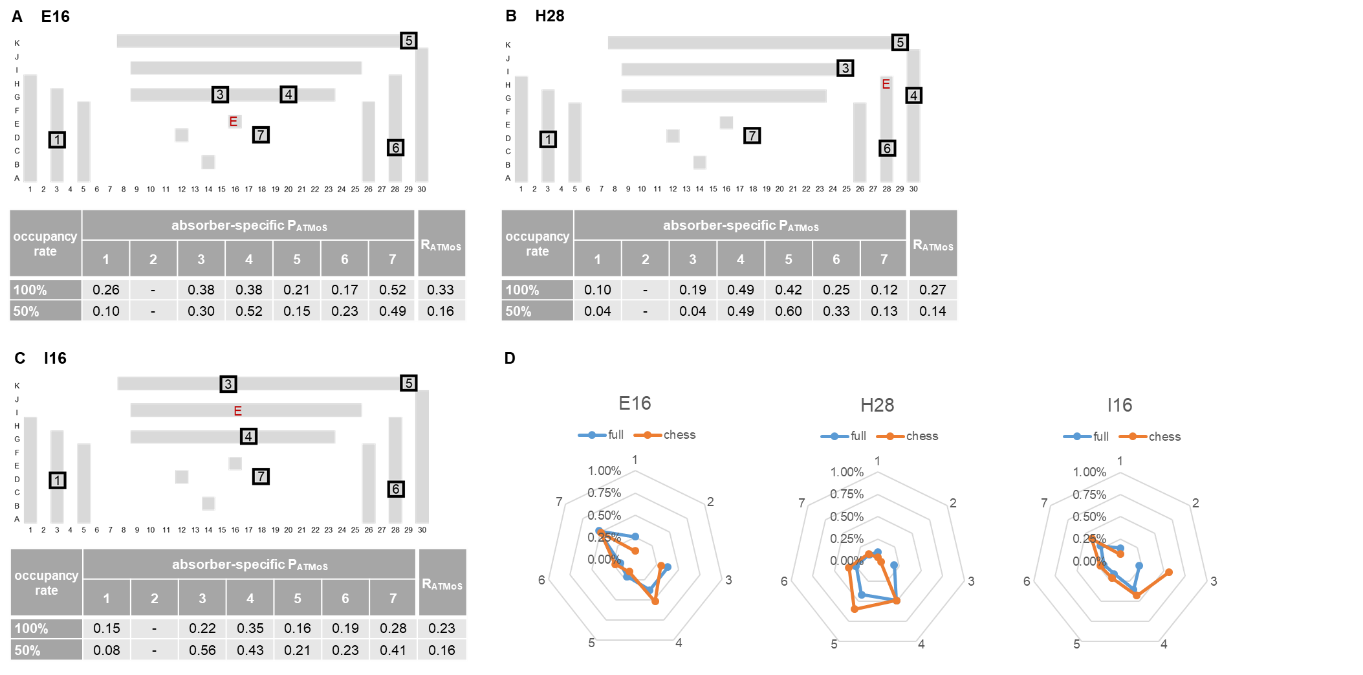

**Fig. S7:	Influence of the checkerboard seating arrangement on experimentally derived individual (P_ATMoS_) and total risk of infection (R_ATMoS_) at MVV1**

The measurement positions for the experimentally derived individual infection risks P_ATMoS_ are shown as black boxes for the emitter (‘E’) positions E16 (A), H28 (B) and I16 (C) for the full occupancy and checkerboard seating arrangement for MVV1. P_ATMoS_ was calculated for each absorber using the absorbed NaCl mass and a quanta emission rate of 18.6 quanta h-1. The values for each absorber are summarised in the table. To obtain R_ATMoS_ the mean value of the seven absorber-specific P_ATMoS_ values of one measurement was calculated and multiplied by the total number of spectators. (D) P_ATMoS_ values for the seven absorbers are summarised in radar charts for each emitter position and occupancy configuration. The numbers 1 to 7 refer to the seven absorbers. The blue solid line showed the P_ATMoS_ values for full occupancy, the orange solid line represents the P_ATMoS_ values for the checkerboard pattern seating.

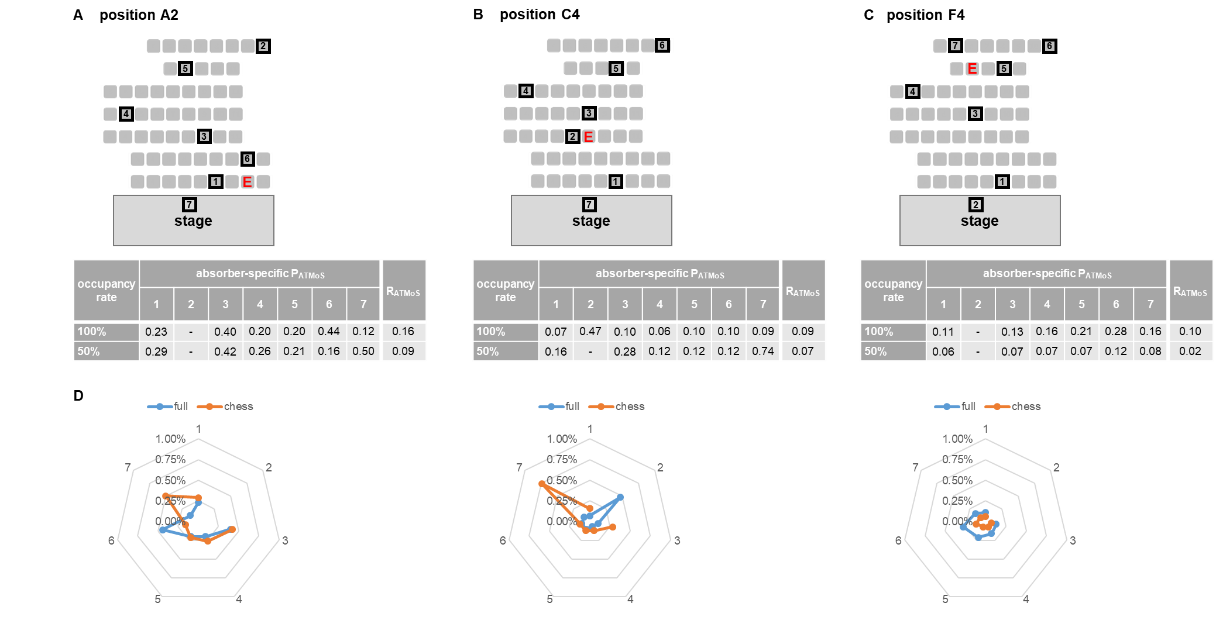

**Fig. S8:	Influence of the checkerboard seating arrangement on experimentally derived individual (P_ATMoS_) and total risk of infection (R_ATMoS_) at MVV2**

The measurement positions for the experimentally derived individual infection risks P_ATMoS_ are shown as black boxes for the emitter (‘E’) positions A2 (A), C4 (B) and F4 (C) for the full occupancy and checkerboard seating arrangement for MVV2. P_ATMoS_ was calculated for each absorber using the absorbed NaCl mass and a quanta emission rate of 18.6 quanta h-1. The values for each absorber are summarised in the table. To obtain R_ATMoS_ the mean value of the seven absorber-specific P_ATMoS_ values of one measurement was calculated and multiplied by the total number of spectators. (D) P_ATMoS_ values for the seven absorbers are summarised in radar charts for each emitter position and occupancy configuration. The numbers 1 to 7 refer to the seven absorbers. The blue solid line showed the P_ATMoS_ values for full occupancy, the orange solid line represents the P_ATMoS_ values for the checkerboard pattern seating.

**Table S1: Room characteristics of the investigated venues**

Experimental measurement time and room characteristics of venues with displacement ventilation DVV1 and DVV2, with mixing ventilation MVV1 and MVV2, and the naturally ventilated venue NVV are listed. The introduction of the ventilation effectiveness E_z_ followed a similar approach of Sun & Zhai (2020) and was adopted from ASHRAE Standard 62.1 (2022, p.22). ^#^Ventilation rate assumption for CFD analysis

| **Venue** | **Number of seats** | **Layout of auditorium** | **Volume of space [m³]** | **Room height [m]** | **Air flow rate**  **[m³/h]** | **Ventilation effectiveness (E_z_)** | **ATMoS measurement time [min]** |
| --- | --- | --- | --- | --- | --- | --- | --- |
| **DVV1** | 99 | ascending | 530 | 3.9 | 4500 | 1.05 | 60 |
| **DVV2** | 470 | ascending | 2300 | 10 | 15.000 | 1.5 | 57 |
| **MVV1** | 103 | ascending | 400 | 3.3 | 6400 | 0.8 | 45 |
| **MVV2** | 60 | ascending | 650 | 5 | 3000 | 0.8 | 45 |
| **NVV** | 244 | ascending | 5000 | 10 | 1500^#^ | 0.5 | 27 |

**Table S2:	Parameter values for the computation of venue-specific risk tables**

| **Parameter** | **Value** | **Unit** |
| --- | --- | --- |
| **Basic quanta emission rate silent, seating E_P0_** | 18.6 | quanta h^-1^ |
| **breathing rate B_0_**  **(susceptibles: silent, seating)** | 0.486 | m³/h |
| **relative quanta emission rate factor (r_E_)** |  |  |
| Silent | 1 |  |
| singing/ shouting | 30 |  |
| **relative breathing rate factor (r_E_)** |  |  |
| Silent | 1 |  |
| singing/ shouting | 1 |  |
| **quanta enhancement due to variants** | 1 |  |
| Alpha | 1.5 |  |
| Delta | 2 |  |
| Omicron | 3.3 |  |
| **face coverings** |  |  |
| no (exhalation, inhalation) | 1 |  |
| exhalation filtration efficiency | 50 | % |
| inhalation filtration efficiency | 30 | % |
| **Contact time** |  |  |
| short | 1 | h |
| medium | 2 | h |
| long | 3 | h |
| **Number of Infective people N_I_** | 1 |  |
